# Impact of COVID-19 on the management and outcomes of ureteric stones in the UK: a multicentre retrospective study

**DOI:** 10.1101/2022.06.27.22276955

**Authors:** Matthew H V Byrne, Fanourios Georgiades, Alexander Light, Catherine E Lovegrove, Catherine Dominic, Josephine Rahman, Senthooran Kathiravelupillai, Tobias Klatte, Kasra Saeb-Parsy, Rajeev Kumar, Sarah Howles, Grant D Stewart, Ben Turney, Oliver Wiseman, the COVID Stones Collaborative

**Author notes:** **Corresponding author** Name: Matthew Byrne, Email address, Full Institution address: Oxford University Hospitals NHS Foundation Trust, UK, Institution post code: OX3 7LE, Twitter: @mhvbyrne. Collaborating authors are listed in Appendix A. **Authorship:** MHVB and FG were responsible for conceptualisation. All authors were responsible for writing the first draft and revisions. AL, CD, JR, RK, and Members of the COVID Stone Collaborative were responsible for data collection. MHVB was responsible for data analysis. SH, GDS, BT, and OW were responsible for supervision. BT and OW are the guarantors. All authors have seen and approved the final version. **Ethical Approval:** We followed NHS Health Research Authority guidance and the study did not require ethical approval. Each participating site obtained local audit approval.

## Abstract

**Objectives:** To determine if management of ureteric stones in the United Kingdom changed during the COVID-19 pandemic and whether this affected patient outcomes.

**Patients and methods:** We conducted a multicentre retrospective study of adults with CT-proven ureteric stone disease at 39 UK hospitals during a pre-pandemic period (23/3/19 to 22/6/19) and a period during the pandemic (the 3-month period after the first SARS-CoV-2 case at individual sites). The primary outcome was success of primary treatment modality, defined as no further treatment required for the index ureteric stone. Our study protocol was published prior to data collection.

**Results:** A total of 3735 patients were included (pre-pandemic=1956 patients; pandemic=1779 patients). Stone size was similar between groups (p>0.05). During the pandemic, patients had lower hospital admission rates (pre-pandemic=54.0% vs pandemic=46.5%, p<0.001), shorter length of stay (mean=4.1 vs. 3.3 days, p=0.02), and higher rates of use of medical expulsive therapy (17.4% vs. 25.4%, p<0.001). In patients who received interventional management (pre-pandemic n=787 vs. pandemic n=685), rates of ESWL (22.7% vs. 34.1%, p<0.001) and nephrostomy were higher (7.1% vs. 10.5%, p=0.03); and rates of ureteroscopy (57.2% vs. 47.5%, p<0.001), stent insertion (68.4% vs. 54.6%, p<0.001), and general anaesthetic (92.2% vs. 76.2%, p<0.001) were lower.

There was no difference in success of primary treatment modality between patient cohorts (pre-pandemic=73.8% vs. pandemic=76.1%, P=0.11), nor when patients were stratified by treatment modality or stone size. Rates of operative complications, 30-day mortality, and readmission and renal function at 6 months did not differ between the data collection periods.

**Conclusions:** During the COVID-19 pandemic, there were lower admission rates and fewer invasive procedures performed. Despite this, there were no differences in treatment success or outcomes. Our findings indicate that clinicians can safely adopt management strategies developed during the pandemic to treat more patients conservatively and in the community.

## INTRODUCTION

Nephrolithiasis is a major clinical and economic health challenge. Up to 20% of men and 10% of women are affected by stone disease ^1,2^. It is responsible for over 85,000 hospital episodes in the UK and costs the UK National Health Service (NHS) an estimated £190-324 million per year ^3^.

The management of ureteric stones can be conservative, as most stones smaller than 5mm pass spontaneously ^4^. However, interventions such as extracorporeal shockwave lithotripsy (ESWL) or ureteroscopic (URS) laser lithotripsy may be required. Untreated stone disease can result in refractory pain, sepsis, renal failure, and death ^5^. The UK National Institute for Health and Care Excellence (NICE) guidelines published in 2019 recommend that adults with ureteric stones measuring less than 10mm should be treated with ESWL and URS is recommended as a second line alternative. For ureteric stones measuring between 10 and 20mm, URS should be offered as first line treatment, and ESWL can be considered if local facilities will allow stone clearance to be achieved within 4 weeks ^6^.

In March 2020, COVID-19 was declared a pandemic by the World Health Organisation (WHO), and measures were introduced across the world to mitigate the spread of infection ^7^. There was worldwide disruption to healthcare provision, including increased pressures on healthcare services and the cancellation of elective procedures ^8^. The multicentre, international COVIDSurg study demonstrated that perioperative infection with the SARS-CoV-2 virus was associated with an unadjusted 30-day mortality of 23.8% ^9^. In light of this, and as a result of reduced access to operating theatres, recommendations were made to favour non-operative management strategies ^8,10–12^. Furthermore, during the peak of the pandemic in the UK, non-COVID-19 related Emergency Department attendances fell dramatically as patients delayed or avoided presentation to hospital due to fear of infection ^13^.

The existing body of literature around COVID-19 and endourology discusses alterations required of clinical care to accommodate the widespread disruption to healthcare services due to COVID-19. However, these articles were written at the start of the COVID-19 pandemic and do not discuss whether these suggested changes to treatment algorithms manifested during the pandemic or what their impact on outcomes were ^11,12,14^.

We hypothesised that during COVID-19 there were delays in patient presentation resulting in higher rates of AKI and sepsis and that non-invasive management options such as observation, ESWL, and alternatives to general anaesthesia were used more frequently, resulting in higher rates of failed index management and subsequent change in treatment modality and/or re-presentation to hospital. We sought to test this hypothesis by undertaking a multicentre, retrospective study to determine how management of ureteric stones changed during the COVID-19 pandemic in the United Kingdom and define how changes in management affected patient outcomes.

## METHODS

### Study design

We conducted a multicentre, retrospective cohort study of the management and outcomes of patients presenting with ureteric stones before and during the COVID-19 pandemic at 39 hospitals in the United Kingdom.

Our protocol was published prior to data collection ^15^. We followed principles of the trainee-led collaborative research model ^16^, coordinated by the COVID Stones Collaborative; and the STROBE (STrengthening the Reporting of OBservational studies in Epidemiology) guidelines ^17^ (Appendix B). NHS Health Research Authority guidance was followed, and each participating site obtained local audit approval.

Patients aged ≥18 years with ureteric stone disease confirmed on contrast or non-contrast computed tomography (CT) imaging were identified via retrospective review of all abdominal CT scans undertaken during relevant data collection periods. Patients with non-ureteric stone disease were excluded.

### Data collection

Data was collected during two time periods: a pre-pandemic period from 23/3/19 to 22/6/19; and a period during the pandemic, which was defined as the 3-month period after the first SARS-CoV-2 case at each individual site. This time point was approximately equivalent to the start of the first UK lockdown due to Covid-19 on 23/3/20. Data was collected by local collaborators, and entered and stored on the Research Electronic Data Capture (REDCap) server managed and hosted by the University of Oxford, UK ^18,19^. Data collected included demographics, management and outcomes at 6 months follow-up.

### Outcomes

Our primary objective was to assess success of primary treatment modality, defined as no additional treatment required for the index ureteric stone. Our secondary objectives were to assess rates of non-operative management, ESWL, stent insertion, ureteroscopy, and nephrostomy insertion; type of anaesthesia for operative management options; operative complications; hospital admission and length of stay; 30-day and 6-month mortality; readmission, and impact of stone on renal function.

### Statistical analysis

Statistical analysis was performed using R (version 4.1.0) ^20^. Patients in the pre-pandemic cohort were compared to patients in the pandemic cohort. Two-sided unpaired t-tests, and Chi-squared tests were used to analyse the data. Data is presented as mean ± standard deviation (SD) or as raw number and percentage. A p-value <0.05 was deemed significant.

## RESULTS

### Demographics

Data were collected from a total of 3735 patients from 39 centres. In the pre-pandemic period data were entered for 1956 patients, and in pandemic period data was entered for 1779 patients. Baseline patient characteristics were broadly similar (Table 1). Although, there was a significant difference in age between cohorts, the median age group for both cohorts was 50-59 years.

**Table 1:** baseline characteristics between the pre-pandemic patient cohort and the pandemic patient cohort. AKI = acute kidney injury CFU = colony forming units; CRP = C reactive protein; IQR = interquartile range; SD = standard deviation; WCC = white cell count.

|  | Pre-pandemic<br>period<br>(N=1956) | Pandemic<br>period<br>(N=1779) | p-value |
| --- | --- | --- | --- |
| **Age group** |  |  | 0.02 |
| 18-19 years | 11 (0.6%) | 17 (1.0%) |  |
| 20-29 years | 198 (10.1%) | 164 (9.2%) |  |
| 30-39 years | 340 (17.4%) | 346 (19.4%) |  |
| 40-49 years | 361 (18.5%) | 359 (20.2%) |  |
| 50-59 years | 409 (20.9%) | 405 (22.8%) |  |
| 60-69 years | 336 (17.2%) | 267 (15.0%) |  |
| 70-79 years | 210 (10.7%) | 149 (8.4%) |  |
| 80+ years | 90 (4.6%) | 72 (4.1%) |  |
| **Gender** |  |  | 0.823 |
| Female | 576 (29.4%) | 517 (29.1%) |  |
| Male | 1380 (70.6%) | 1262 (70.9%) |  |
| **BMI** |  |  |  |
| Mean (SD) | 28.8 (6.3) | 29.6 (7.4) | 0.074 |
| Median [IQR] | 28.0 [24.4-32.1] | 28.7 [24.7-32.9] |  |
| **Non-urolological comorbidities** |  |  | 0.075 |
| Yes | 538 (36.2%) | 453 (32.9%) |  |
| No | 950 (63.8%) | 923 (67.1%) |  |
| **Urological comorbidities** |  |  | 0.662 |
| Yes | 56 (3.8%) | 46 (3.4%) |  |
| No | 1429 (96.2%) | 1309 (96.6%) |  |
| **Ureteric stone location?** |  |  | 0.52 |
| Left | 1014 (51.8%) | 929 (52.2%) |  |
| Right | 873 (44.6%) | 799 (44.9%) |  |
| Bilateral | 69 (3.53%) | 51 (2.87%) |  |
| **Maximum stone size (mm)** |  |  | 0.687 |
| ≤5 | 1026 (57.5%) | 926 (56.9%) |  |
| 6-10 | 633 (35.5%) | 598 (36.8%) |  |
| 11-19 | 108 (6.1%) | 85 (5.2%) |  |
| ≥20 | 18 (1.0%) | 17 (1.1%) |  |
| **Active infection at time of presentation** |  |  | 0.032 |
| Yes | 258 (13.2%) | 193 (10.8%) |  |
| No | 1697 (86.8%) | 1586 (89.2%) |  |
| **Fever (Temperature >38 degree C) at any stage in index admission** |  |  | 0.031 |
| Yes | 151 (9.3%) | 107 (7.1%) |  |
| No | 1482 (90.8%) | 1406 (92.9%) |  |
| **Admission blood tests CRP (mg/L)** |  |  | 0.562 |
| Mean (SD) | 27.7 (64.6) | 26.4 (68.7) |  |
| Median [IQR] | 4.0 [1.0-17.0] | 3.0 [1.0-13.0] |  |
| **Admission blood tests WCC (x10<sup>9</sup>/L)** |  |  | 0.725 |
| Mean (SD) | 11.0 (6.2) | 11.0 (4.2) |  |
| Median [IQR] | 10.3 [8.1-12.9] | 10.7 [8.1-13.1] |  |
| **Positive blood culture** |  |  | 0.749 |
| Yes | 51 (38.1%) | 41 (41.0%) |  |
| No | 83 (61.9%) | 59 (59.0%) |  |
| **Positive urine culture (>10<sup>5</sup> cfu)** |  |  | 0.796 |
| Yes | 97 (21.8%) | 78 (20.9%) |  |
| No | 347 (78.2%) | 296 (79.1%) |  |
| **Admission blood tests Creatinine (umol/L)** |  |  | 0.765 |
| Mean (SD) | 105 (70.8) | 106 (75.9) |  |
| Median [IQR] | 94.0 [77.0-113.0] | 93.0 [78.0-113.0] |  |
| **AKI** |  |  | 0.041 |
| Yes | 340 (21.9%) | 290 (18.9%) |  |
| No | 1211 (78.1%) | 1245 (81.1%) |  |
| **AKI Grade (KGIDO)** |  |  |  |
| Grade 1 | 226 (74.1%) | 202 (74.8%) |  |
| Grade 2 | 55 (18.0%) | 44 (16.3%) |  |
| Grade 3 | 24 (7.9%) | 24 (8.9%) |  |

Stone site and size were similar between cohorts (Table 1). Unexpectedly, patients from the pre-pandemic cohort were reported to have significantly higher rates of active infection and acute kidney injury on admission, and fever at any point during index admission. However, mean C-reactive protein, white cell count, positive microbiology cultures (urine or blood), and creatinine were not significantly different (Table 1).

### Management

Overall, patients in the pandemic period had significantly lower rates of admission to hospital (pre-pandemic 54.0% vs. pandemic 46.5%, p<0.001) and shorter length of stay in hospital (mean length of stay in days, pre-pandemic=4.1 (SD=8.0) vs. pandemic=3.3 (SD=5.9), p=0.02) compared to the pre-pandemic period. The use of alpha blockers was significantly higher during the pandemic (pre-pandemic=17.4% vs. pandemic=25.4%, p<0.001). Despite the higher rates of active infection and fever, antibiotic usage was similar between cohorts (Table 2).

**Table 2:** Management and complications

|  | Pre-pandemic period<br>(N=1956) | Pandemic period<br>(N=1779) | P-value |
| --- | --- | --- | --- |
| **Admitted to hospital?** |  |  | <0.001 |
| Yes | 1057 (54.0%) | 827 (46.5%) |  |
| No | 899 (46.0%) | 952 (53.5%) |  |
| **Length of stay (days)** |  |  | 0.021 |
| Mean (SD) | 4.08 (8.04) | 3.29 (5.94) |  |
| **Alpha blocker** |  |  | <0.001 |
| Yes | 316 (17.4%) | 414 (25.4%) |  |
| No | 1501 (82.6%) | 1219 (74.6%) |  |
| **Antibiotics** |  |  |  |
| Yes-Oral | 171 (8.74%) | 148 (8.32%) | 0.251 |
| Yes-IV | 304 (15.5%) | 245 (13.8%) |  |
| No | 1481 (75.7%) | 1386 (77.9%) |  |
| **Interventional management** |  |  | 0.295 |
| Yes | 787 (40.2%) | 685 (38.5%) |  |
| No | 1169 (59.8%) | 1094 (61.5%) |  |
|  | Pre-pandemic period<br>(N=787) | Pandemic period<br>(N=685) | P-value |
| **Anaesthetic** |  |  | <0.001 |
| General | 390 (92.2%) | 173 (76.2%) |  |
| Non-general | 33 (7.80%) | 54 (23.8%) |  |
| **ESWL for index stone** |  |  | <0.001 |
| Yes | 179 (22.7%) | 233 (34.1%) |  |
| No | 608 (77.3%) | 451 (65.9%) |  |
| **Ureteroscopy for index stone** |  |  | <0.001 |
| Yes | 450 (57.2%) | 325 (47.5%) |  |
| No | 337 (42.8%) | 359 (52.5%) |  |
| **Stent insertion for index stone (alone or as part of Ureteroscopy)** |  |  | <0.001 |
| Yes | 538 (68.4%) | 374 (54.6%) |  |
| No | 249 (31.6%) | 311 (45.4%) |  |
| **Emergency nephrostomy insertion for index stone** |  |  | 0.027 |
| Yes | 56 (7.1%) | 72 (10.5%) |  |
| No | 731 (92.9%) | 613 (89.5%) |  |
| **Operative complication** |  |  | 0.616 |
| Yes | 87 (11.5%) | 68 (10.5%) |  |
| No | 671 (88.5%) | 580 (89.5%) |  |
| 30-day highest Clavien-Dindo grade |  |  |  |
| None | 671 (88.5%) | 580 (89.5%) |  |
| I | 44 (5.8%) | 28 (4.3%) |  |
| II | 29 (3.8%) | 15 (2.3%) |  |
| III-A | 1 (0.1%) | 8 (1.2%) |  |
| III-B | 9 (1.2%) | 12 (1.9%) |  |
| IV-A | 2 (0.3%) | 0 (0.0%) |  |
| IV-B | 1 (0.1%) | 2 (0.3%) |  |
| V (death) | 1 (0.1%) | 3 (0.5%) |  |

#### Interventional management

There were no differences in rates of interventional management between the cohorts (pre-pandemic, n=787 [40.2%] vs. pandemic, n=685 [38.5%], p=0.30), or ASA grade (p=0.50) and WHO/ECOG Performance status (p=0.21) in patients who received operative management (Table S1). However, there were significantly higher rates of ESWL and nephrostomy insertion, and significantly lower rates of general anaesthetic, ureteroscopy and stent insertion during the pandemic compared to pre-pandemic (Figure 1, Table 2). During the pandemic, planned interventions were delayed due to COVID-19 status in 63 out of 1580 patients (4.0%).

**Figure 1:**
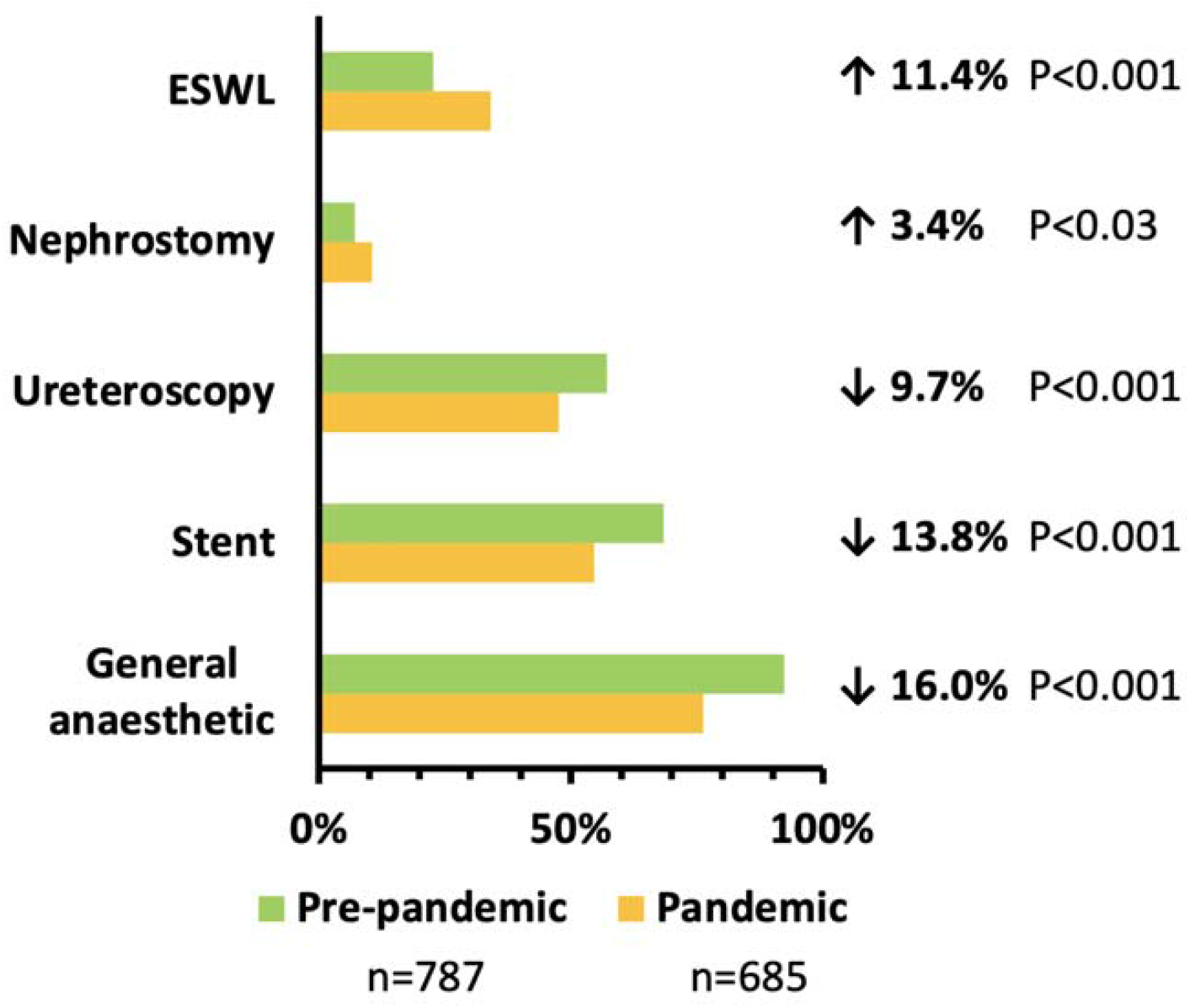
Change in interventional management during the pandemic

### Outcomes

#### 30-day outcomes

The 30-day operative complication rate among those who received operative management was similar between the pre-pandemic and pandemic cohort (Table 2), as was the 30-day mortality rate (any cause) across the whole of each cohort (pre-pandemic=8 of 1793 [0.5%] vs. pandemic=9 of 1640 [0.6%], p=0.85).

#### 6-month outcomes

The success rate of primary treatment (i.e. no further treatment required for the index stone after primary treatment modality) was similar between cohorts (pre-pandemic=73.8% vs. pandemic=76.1%, p=0.11). There was no significant difference in success of primary treatment modality when stratified by treatment modality or stone size.

In patients who did require further intervention, the rates of ESWL, ureteroscopy, retrograde stent insertion, and nephrostomy were similar between each cohort (Table 3).

**Table 3:**
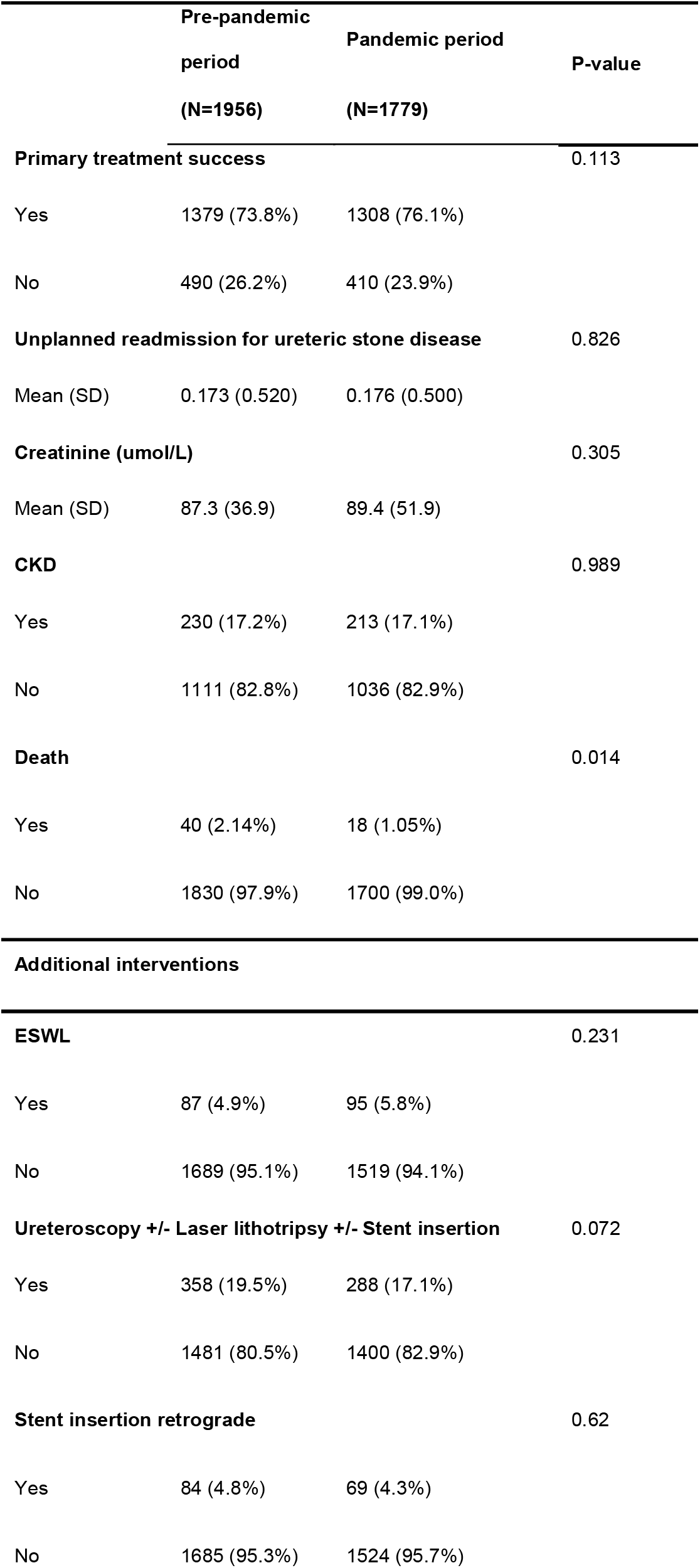
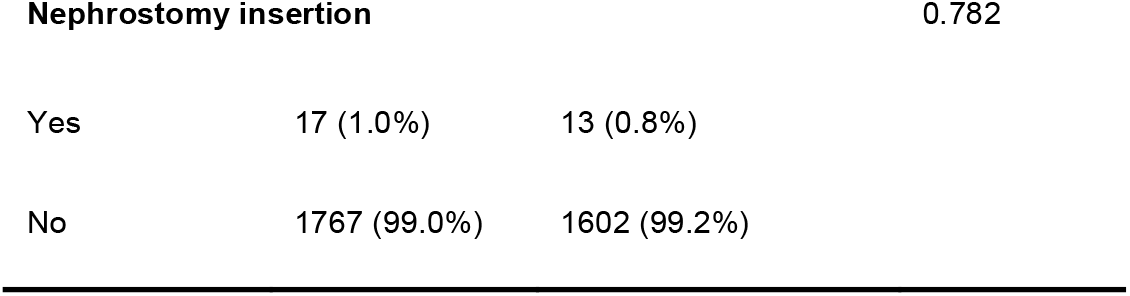
Outcomes and additional operative management required at 6 months follow-up.

|  | Pre-pandemic<br>period<br>(N=1956) | Pandemic period<br>(N=1779) | P-value |
| --- | --- | --- | --- |
| **Primary treatment success** |  |  | 0.113 |
| Yes | 1379 (73.8%) | 1308 (76.1%) |  |
| No | 490 (26.2%) | 410 (23.9%) |  |
| **Unplanned readmission for ureteric stone disease** |  |  | 0.826 |
| Mean (SD) | 0.173 (0.520) | 0.176 (0.500) |  |
| **Creatinine (umol/L)** |  |  | 0.305 |
| Mean (SD) | 87.3 (36.9) | 89.4 (51.9) |  |
| **CKD** |  |  | 0.989 |
| Yes | 230 (17.2%) | 213 (17.1%) |  |
| No | 1111 (82.8%) | 1036 (82.9%) |  |
| **Death** |  |  | 0.014 |
| Yes | 40 (2.14%) | 18 (1.05%) |  |
| No | 1830 (97.9%) | 1700 (99.0%) |  |
| **Additional interventions** |  |  |  |
| **ESWL** |  |  | 0.231 |
| Yes | 87 (4.9%) | 95 (5.8%) |  |
| No | 1689 (95.1%) | 1519 (94.1%) |  |
| **Ureteroscopy +/- Laser lithotripsy +/- Stent insertion** |  |  | 0.072 |
| Yes | 358 (19.5%) | 288 (17.1%) |  |
| No | 1481 (80.5%) | 1400 (82.9%) |  |
| **Stent insertion retrograde** |  |  | 0.62 |
| Yes | 84 (4.8%) | 69 (4.3%) |  |
| No | 1685 (95.3%) | 1524 (95.7%) |  |

**Nephrostomy insertion** 0.782
|  |  |  |
| --- | --- | --- |
| Yes | 17 (1.0%) | 13 (0.8%) |
| No | 1767 (99.0%) | 1602 (99.2%) |

The mean number of unplanned admissions (pre-pandemic=0.17 (SD=0.52) vs. pandemic=0.18 (SD=0.50), p=0.83), mean creatinine (pre-pandemic=87.3 (SD=36.9) vs. pandemic=89.4 (SD=51.9) μmol/L, p=0.31), and chronic kidney disease stage 1 to 5 (pre-pandemic=17.1% vs. pandemic=17.9%, p=0.99) were similar between cohorts. There was a higher mortality rate in the pre-pandemic cohort group (pre-pandemic=40 of 1830 [2.1%] vs. pandemic=18 of 1700 [1.1%], p=0.01).

## DISCUSSION

In response to the COVID-19 pandemic, strategies to prioritise and triage patients with urological pathology were developed ^12^. These included conservative treatment whenever possible ^12^, and use of local-anaesthesia to minimise ventilator use and reduce risk of COVID-19 exposure ^11,12,14^. If operative management was deemed necessary, recommendations were made to select patients according to surgical priority using patient factors (symptoms, comorbidities, and renal tract abnormalities) and stone factors (obstruction, infection, and conservative management failure) ^11,12^. In cases where there was an infected, obstructed system, multiple sources recommended insertion of a ureteric stent under local anaesthetic as first line treatment, with nephrostomy as second line option ^12,14^.

In this study, we demonstrate that the management of ureteric stones changed across the UK during at the onset of the COVID-19 pandemic, with fewer invasive procedures and increased rates of ESWL and medical expulsive management. We also found that rates of nephrostomy were higher, despite lower rates of AKI and active infection.

Other studies have reported a decrease in urological presentations during the COVID-19 pandemic ^21–24^ We therefore predicted that during the pandemic patients with ureteric colic would have delayed presenting to hospital and therefore have been a more unwell cohort. However, in our data, rates of AKI and active infection were lower in the pandemic patient cohort suggesting that these patients did not delay their presentation long enough to impact their clinical condition. These findings contrast with other urolithiasis datasets. Castellani et al. compared ureteric stone disease outcomes of 298 patients prior to the pandemic with 218 patients during the pandemic, reporting reduced admissions and higher rates of infected-obstructed systems, hospitalisation, and intervention during the COVID-19 pandemic^25^. Furthermore, Flammia et al. found that serum creatinine was significantly higher in 36 patients with urinary stone emergencies during the pandemic compared to 44 patients with urinary stone emergencies prior to the pandemic, which the authors posited was due to delayed presentation ^26^. Similarly, Gul et al. demonstrated an increase in creatinine, white cell count, hospital admissions, antibiotic treatment, and emergency nephrostomy insertion in 35 patients with urolithiasis during the pandemic compared to 114 patients with urolithiasis prior to the pandemic ^27^.

Our findings are consistent with those reported by Anderson et al. and Nourian et al. who identified no difference in markers of infection or AKI between pre-pandemic and post-pandemic cohorts of patients presenting with urolithiasis ^28, 29^. Our study represents the largest cohort to date investigating ureteric stone outcomes during the pandemic, has a multicentre design and had a pre-defined protocol. Thus, we predict our findings are representative of true outcomes in the UK during the COVID-19 pandemic.

Steinberg et al. suggested that we can use the COVID-19 pandemic as an opportunity to reassess ureteric stone management strategies, and establish whether conservative management strategies have been under-used ^30^. Our study is the first to evaluate the impact of changes in the management of ureteric stones during the COVID-19 pandemic on patient outcomes at 6 month follow up ^26,28,29,31,32^. We demonstrate that increased use of conservative management strategies did not have a detrimental effect on primary treatment success or patient outcomes at 6-month follow up. Our data supports increased use of less-invasive options recommended by NICE guidance including watchful waiting with medical expulsive therapy and ESWL as first line – there was an approximately 10% shift from URS to ESWL during the pandemic with non-inferior outcomes. Our data also support a reduction in admission rates and earlier discharge. It is unclear whether these changes will revert once the pandemic stabilises and patient backlog is tackled or if practice will change permanently. Evidence such as this should drive a more permanent change to less-invasive management, as the change in practice during the pandemic has shown that this is safe and as effective.

This study is the first of its kind to be conducted across multiple sites with 6-month follow-up. However, the study is limited by its retrospective design, and missing data. Our study was conducted across 39 centres and the number of cases entered by each centre varied considerably. This may be due to differences in local patient populations, however, it may be that not all patients within each time period were captured. This increases the risk of selection bias within our study and therefore no comment can be made on whether there was a change in the number of presentations with ureteric stone disease during each time period.

## CONCLUSIONS

As a result of the COVID-19 pandemic patients in the UK with ureteric stones were less likely to be treated invasively and more likely to be managed without admission. However, this change in practice did not result in inferior outcomes for patients. The pandemic has given the urological community an opportunity to re-evaluate management of ureteric stones; our findings indicate that a greater proportion of ureteric stones can be managed safely and effectively with non-invasive ambulatory options.

## Data Availability

MHVB and FG were responsible for conceptualisation. All authors were responsible for writing the first draft and revisions. AL, CD, JR, RK, and Members of the COVID Stone Collaborative were responsible for data collection. MHVB was responsible for data analysis. SH, GDS, BT, and OW were responsible for supervision. BT and OW are the guarantors. All authors have seen and approved the final version.

## Acknowledgements

None

## SUPPLEMENTARY TABLES

**Table S1:** ASA Grade and WHO/ECOG Performance status

|  | Pre-pandemic period<br>(N=787) | Pandemic period<br>(N=685) | P-value |
| --- | --- | --- | --- |
| **ASA grade** |  |  |  |
| 1. A normal healthy patient | 219 (42.0%) | 134 (43.4%) | 0.498 |
| 2. A patient with mild systemic disease | 199 (38.1%) | 125 (40.5%) |  |
| 3. A patient with severe systemic disease | 89 (17.0%) | 45 (14.6%) |  |
| 4. A patient with severe systemic disease that is a constant threat to life | 15 (2.87%) | 5 (1.62%) |  |
| **WHO/ECOG Performance status** |  |  |  |
| Fully active, able to carry on all pre-disease activities | 326 (73.1%) | 211 (75.9%) | 0.214 |
| Restricted in physical strenuous activity, able to perform light work | 87 (19.5%) | 39 (14.0%) |  |
| Ambulatory and capable of all self-care, unable to work | 18 (4.04%) | 15 (5.40%) |  |
| Capable of limited self-care only | 11 (2.47%) | 7 (2.52%) |  |
| Completely disabled. Confined to bed or chair | 4 (0.897%) | 6 (2.16%) |  |

